# Symptomatic SARS-CoV-2 re-infection of a health care worker in a Belgian nosocomial outbreak despite primary neutralizing antibody response

**DOI:** 10.1101/2020.11.05.20225052

**Authors:** Philippe Selhorst, Sabrina Van Ierssel, Jo Michiels, Joachim Mariën, Koen Bartholomeeusen, Eveline Dirinck, Sarah Vandamme, Hilde Jansens, Kevin K. Ariën

**Author notes:** corresponding author Nationalestraat 155 2000 Antwerpen BELGIUM.

## Abstract

**Background:** It is currently unclear whether SARS-CoV-2 re-infection will remain a rare event, only occurring in individuals who fail to mount an effective immune response, or whether it will occur more frequently when humoral immunity wanes following primary infection.

**Methods:** A case of re-infection was observed in a Belgian nosocomial outbreak involving 3 patients and 2 health care workers. To distinguish re-infection from persistent infection and detect potential transmission clusters, whole genome sequencing was performed on nasopharyngeal swabs of all individuals including the re-infection case’s first episode. IgA, IgM, and IgG and neutralizing antibody responses were quantified in serum of all individuals, and viral infectiousness was measured in the swabs of the reinfection case.

**Results:** Re-infection was confirmed in a young, immunocompetent health care worker as viral genomes derived from the first and second episode belonged to different SARS-CoV-2 clades. The symptomatic re-infection occurred after an interval of 185 days, despite the development of an effective humoral immune response following symptomatic primary infection. The second episode, however, was milder and characterized by a fast rise in serum IgG and neutralizing antibodies. Although contact tracing and virus culture remained inconclusive, the health care worker formed a transmission cluster with 3 patients and showed evidence of virus replication but not of neutralizing antibodies in her nasopharyngeal swabs.

**Conclusion:** If this case is representative of most Covid-19 patients, long-lived protective immunity against SARS-CoV-2 might not be likely.

## Introduction

The mechanism, extent, and duration by which primary SARS-CoV-2 infection provides immunity against re-infection are currently unclear. For common cold coronaviruses, loss of immunity and re-infection with the same virus have been reported to occur frequently 12 months after primary infection [1]. For SARS-CoV-2, persistent viral shedding can occur over prolonged periods of time following clinical recovery [2]. To confirm genuine re-infection, whole-genome sequencing is required. Since the start of the SARS-CoV-2 pandemic 11 months ago, only 8 published and confirmed cases of re-infection were reported in Hong Kong [3], the USA [4,5], Belgium [6], the Netherlands [7], Ecuador [8], and India [9]. These cases likely are an underestimate due to the limited detection of asymptomatic cases. In fact many more cases were reported in the media and on preprint servers [2,10–12]. Hence, it remains to be seen whether these re-infection cases represent the tail end of the distribution with many more to come or whether SARS-CoV-2 re-infection remains a rare event.

One hypothesis could be that re-infections occurs as a result of immune evasion by another variant of SARS-CoV-2. However, the genomic variations seen across SARS-CoV-2 sequences are limited and likely the result of neutral evolution, rather than adaptive selection, and although the D614G mutation in Spike has become consensus, there is no evidence that this mutation is linked to host immune pressure [13,14].

Another hypothesis could be that these re-infections only occur in individuals that do not develop an effective immune response during primary infection. Indeed, not all Covid-19 patients seroconvert [15,16] and not all who seroconvert develop neutralizing antibodies [17]. Furthermore, disease severity seems to correlate with higher IgG [18–20] and neutralizing antibody titers [19,20]. Unfortunately, in only two of all reported re-infection cases antibody testing was reported after the first episode. In the Hong Kong case, IgGs but no neutralizing antibodies were detected 10 and 43 days pso [3,21] whereas in the Ecuador case, IgMs but no IgGs were detected by rapid test 4 days pso [8]. Neutralizing antibodies were not measured. Hence, it is currently unclear whether these re-infection cases were able to mount an effective immune response after primary infection.

Finally, it is not known whether immunity prevents onward transmission from those who are re-infected. Most reported re-infection cases [3,4,6–8,11] showed nasopharyngeal samples with high RT-qPCR Ct values, from which virus is usually unculturable. Five Indian health care workers however, displayed high viral loads during their secondary infection but no viral culture was performed to determine the infectiousness of their virus [9,12].

Here we describe a case of symptomatic re-infection in a health care worker despite having developed a neutralizing antibody response following symptomatic primary infection. The re-infection occurred with an interval of 185 days during a nosocomial outbreak involving 5 individuals. Whole genome sequencing was performed, and humoral immune responses and viral infectiousness were quantified.

## Materials & Methods

### Sample collection and diagnosis

Sample collection and clinical evaluation were performed in view of diagnosis and standard of care and approved by the hospital’s ethical committee (EC/PM/nvb/2020.084). Oral consent was obtained from all patients before sampling followed by written consent prior to publication. Initial SARS-CoV-2 diagnosis was performed at the hospital on nasopharyngeal swabs using the Xpert Xpress SARS-CoV-2 test on the GeneXpert® Platform (Cepheid, USA) as per manufacturer’s instructions or by in-house PCR [22] with extraction on NucliSens EasyMag® (Biomérieux, France) and amplification on Cobas LightCycler® (Roche, Switzerland). Complete blood counts were performed on XN-9100® (Sysmex) and biochemistry parameters on Atellica® (Siemens, Germany).

### RNA extraction and RT-qPCR on nasopharyngeal samples

At the Institute of Tropical Medicine Antwerp, RNA was extracted from UTM or eSwab medium after inactivation at 56°C with proteinase K using a Maxwell RSC Instrument. RNA from phocine distemper virus was added to all samples as an internal extraction and PCR inhibition control [23]. A SARS-CoV-2-specific RT-qPCR was then performed to amplify a 112 bp fragment of the E gene as previously described [22] with 5 µL RNA in a 25 µL reaction using the Bioline SensiFAST mix, Reverse Transcriptase and RiboSafe RNase inhibitor. To determine the presence of SARS-CoV-2 replicase activity, a negative strand RT-qPCR was performed as previously described [24] with FWD-Tag-primer-1 (catacgcacggataaa-GCAAGAGATGGTTGTGTTCCC), Tag-primer-1 (catacgcacggataaa), REV-primer-1 (GTAAATGTTGTACCATCACACG) and FAM-labeled negative-strand-probe-1 (CAGCAGCCAAACTAATGGTTGTCATA).

### Whole genome sequencing using MinION

Whole genome sequencing was performed on an Oxford Nanopore MinION device using R9.4 flow cells (Oxford Nanopore Technologies, UK) after a multiplex PCR with an 800bp SARS-CoV-2 primer scheme as previously described [25]. Sequence reads were basecalled in high accuracy mode and demultiplexed using the Guppy algorithm v3.6. Reads were aligned to the reference genome Wuhan-Hu-1 (MN908947.3) with Burrows-Wheeler Aligner (BWA-MEM) and a majority rule consensus was produced for positions with ≥100x genome coverage, while regions with lower coverage, were masked with N characters.

### Phylogenomic analysis

All SARS-CoV-2 genomes were compared at the nucleotide and amino acid level to the reference genome Wuhan-Hu-1. Clade assignment was performed using NextClade v0.7.2 [26]. BLAST+ was used to extract the top 15 matches for each of our sequences from the msa_0929.fasta file downloaded from GISAID (Global Initiative on Sharing All Influenza Data). In addition, we included the most recent (Aug 16-31, 2020) Belgian sequences and all Belgian L, O, V clade sequences collected between March 1-16, 2020. Sequence alignment was performed using MAFFT v7 and a maximum likelihood phylogenetic tree was inferred with IQ-TREE v1.6.12., using the TIM2+F+I model and 500 nonparametric bootstraps, and visualized in FigTree v1.4.4.

### SARS-CoV-2 specific antibody detection tests

The Elecsys electrochemiluminescence immunoassay on the Cobas 8000® analyzer (Roche Diagnostics, Belgium) was used for the qualitative detection of total antibodies against the Nucleocapsid (N) antigen of SARS-CoV-2. A signal threshold ≥1 was defined as positive. For the separate quantification of IgM, IgG, and IgA antibodies, we used a Luminex bead-based assay [27]. In short, recombinant receptor binding domain (RBD) and N protein (BIOCONNECT, The Netherlands) were coupled to 1.25×10^6^ paramagnetic MAGPLEX COOH-microspheres from Luminex Corporation (Texas, USA). After incubation of beads and diluted sera (1/300 for IgG and IgM, 1/100 for IgA), a biotin-labeled anti-human IgG, IgA, and IgM (1:125) and streptavidin-R-phycoerythrin (1:1000) conjugate was added. Beads were read using a Luminex® 100/200 analyzer with 50 µL acquisition, DD gat 5000 - 25000 settings, and high PMT option. Results were expressed as crude median fluorescent intensities (MFI). Samples were considered positive if MFI >3x SD + mean of negative controls (n=16).

### SARS-CoV-2 viral neutralization test and virus isolation

Serial dilutions (1/50 - 1/1600) of heat-inactivated (30 min at 56°C) serum or nasopharyngeal samples were incubated with 3x TCID100 of a SARS-CoV-2 primary isolate (2019-nCoV-Italy-INMI1) for 1h at 37°C / 7% CO2 and subsequently added to 18.000 Vero cells per well for a further 5 days incubation. Assay medium consisted of EMEM (Lonza, Belgium) supplemented with 2 mM L-glutamine, 2% fetal bovine serum, and penicillin - streptomycin (Lonza). After incubation, cytopathic effect caused by viral growth was scored microscopically. 50% (NT50) or 90% (NT90) neutralization titers were calculated using the Reed-Muench method. Similarly, virus isolation was attempted by incubating a serial dilution of nasopharyngeal samples on VeroE6-TMPRSS2 cells after 2 hours of spinoculation at 2500g and 25°C and following up cytopathic effect.

## Results

### Clinical evolution of the outbreak

In September 2020, a nosocomial outbreak occurred at an internal medicine ward in a Belgian hospital. A man in his seventies (PAT1) developed influenza-like symptoms of cough, low grade fever and general malaise and tested positive for SARS-CoV-2. Three days later a woman, also in her seventies, (PAT2) tested positive after developing gastro-intestinal symptoms and general malaise (Table 1). In response to this outbreak all patients and health care workers on the ward were tested revealing 1 additional asymptomatic man in his eighties (PAT3) and 2 infected health care workers (HCW1 & HCW2) showing mild symptoms. All individuals recovered completely. HCW1, a woman in her thirties, had already been infected with SARS-CoV-2 in March 2020. During this first episode she had a protracted mild illness with cough, dyspnea, headache, fever and general malaise. Her hematological and biochemical parameters were consistent with viral infection showing decreased white blood cell counts and mildly elevated CRP (Supplementary table 1). She was managed as an outpatient and slowly resumed work after 1 month. During the second episode her clinical presentation was milder, and she resumed work 10 days after diagnosis although she experienced dyspneic spells for up to 3 weeks. A blood sample and a second swab were taken 7 days after symptom onset. This time no laboratory abnormalities were found (Supplementary table 1).

**Table 1.**
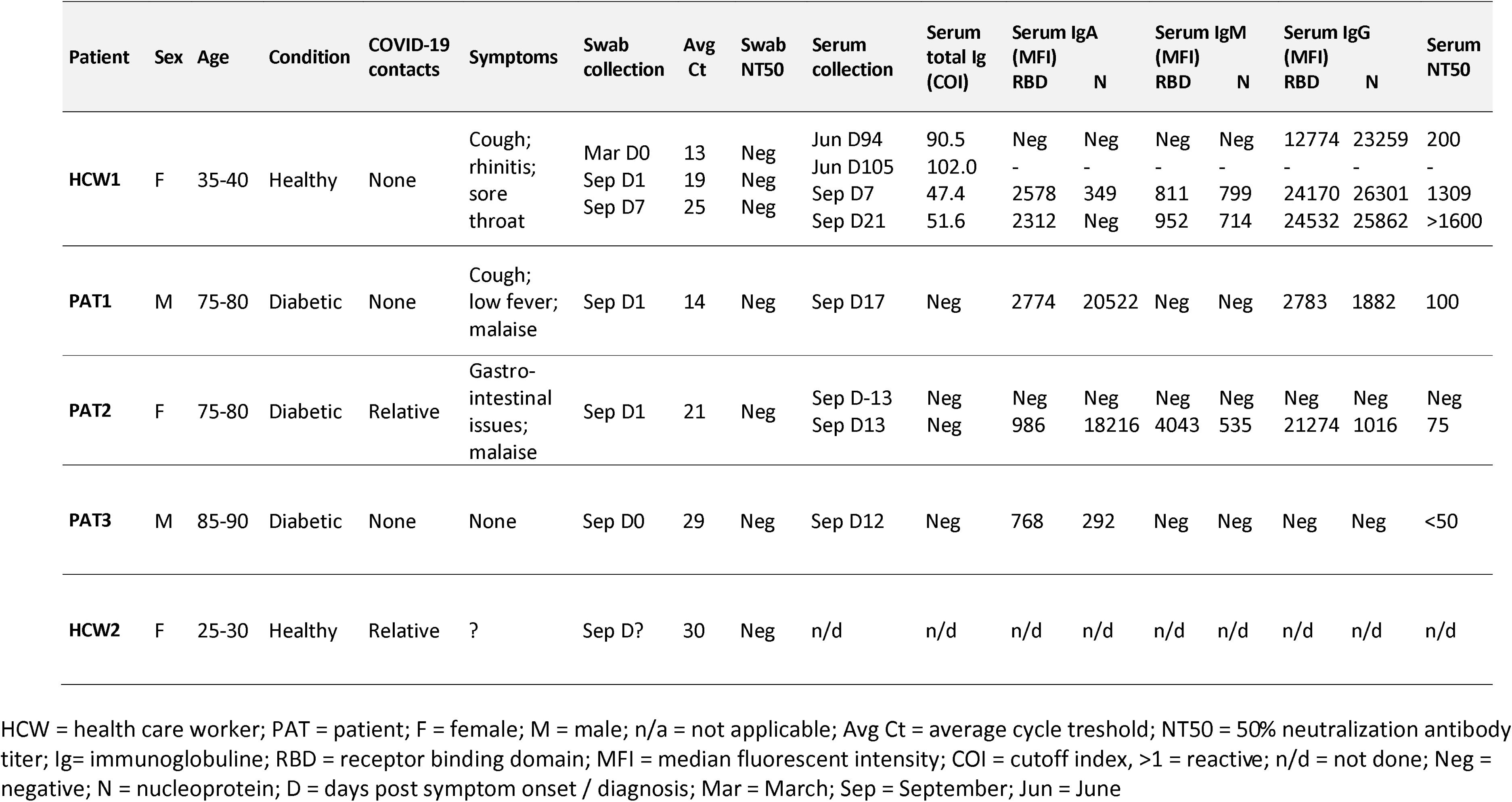
Epidemiological and serological data on the 3 patients and 2 health care workers involved in the nosocomial outbreak.

### Whole genome sequencing revealing transmission cluster

To distinguish re-infection from persistent infection and detect potential transmission clusters, whole genome sequencing was performed on all nasopharyngeal swabs taken from the 3 patients and 2 health care workers, including the swab taken from HCW1 during her first episode. 99.6% of the SARS-CoV-2 genome (nucleotide 55–29823) was recovered at an average mean depth of 771-fold (Supplementary table 2). Analysis of these sequences (EPI_ISL_582127–32) revealed that the virus which infected HCW1 in September belonged to a different SARS-CoV-2 clade (G clade) than the virus causing her first COVID-19 episode in March (V clade) (Figure 1). A total of 18 nucleotide differences, of which only one shared (i.e. G11083T defining the V clade from which clade G emerged), could be observed between the 2 strains (Figure 2). This confirms their distinct nature and is within the range of 9-24 nucleotide differences reported for other re-infections. Interestingly, the 3 infected patients shared the same virus and hence constitute a recent transmission cluster with HCW1. None of the amino acid mutations in the S gene of the reinfecting virus were previously reported to confer resistance to convalescent plasma or RBD-specific monoclonal antibodies [28]. Finally, the viral sequence derived from HCW2 differed by 27 nucleotides from HCW1’s virus and belonged to GISAID’s GH clade (Figure 1 and 2). This suggests that HCW2 was infected by an external transmission source which corresponds with the fact that one of her relatives also tested positive for SARS-CoV-2. As the phylogenetic analysis and visualization provided by Nextstrain [29] only uses a subsample of publicly available SARS-CoV-2 genomes, we performed a blast search to include the most closely related known sequences in our analysis. As expected, the genomes from HCW1, PAT13, and HCW2 in September were closely related to Belgian sequences obtained in July-September whereas the virus from HCW1’s March episode was identical to Belgian sequences collected in that same month (Supplementary figure 1).

**Figure 1.**
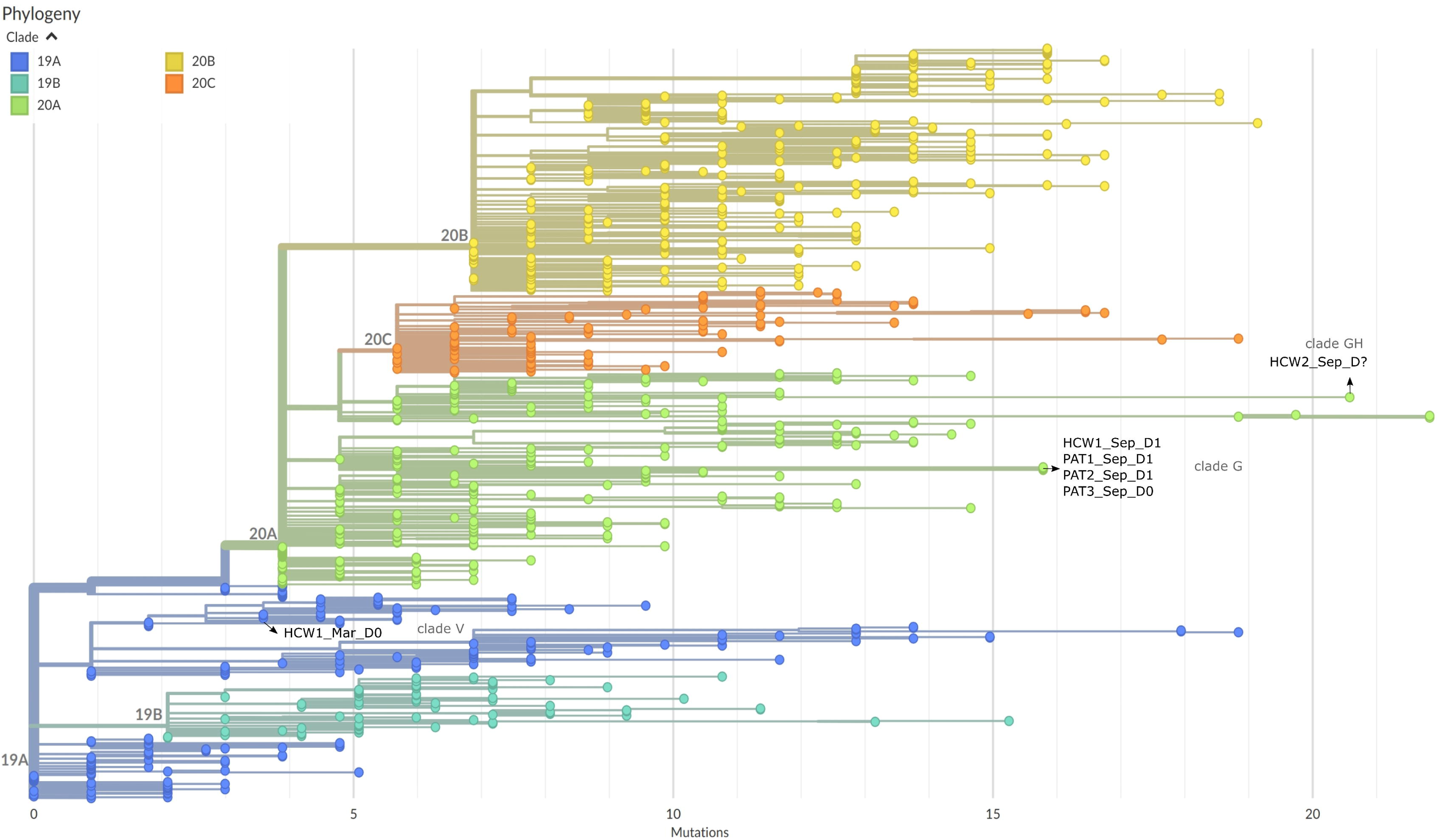
SARS-CoV-2 clade assignment of 6 genomes recovered from 3 patients (PAT) and 2 health care workers (HCW). Sequences were aligned to a representation of the global SARS-CoV-2 genetic diversity using a banded Smith-Waterman algorithm with an affine 160 gap-penalty and subsequently visualized using NextClade v0.7.2. Colors represent the different GSIAID clades. The 6 genomes and their clades are marked by respectively arrows and grey text. D = days post symptom onset / diagnosis; Mar = March; Sep = September

**Figure 2.**
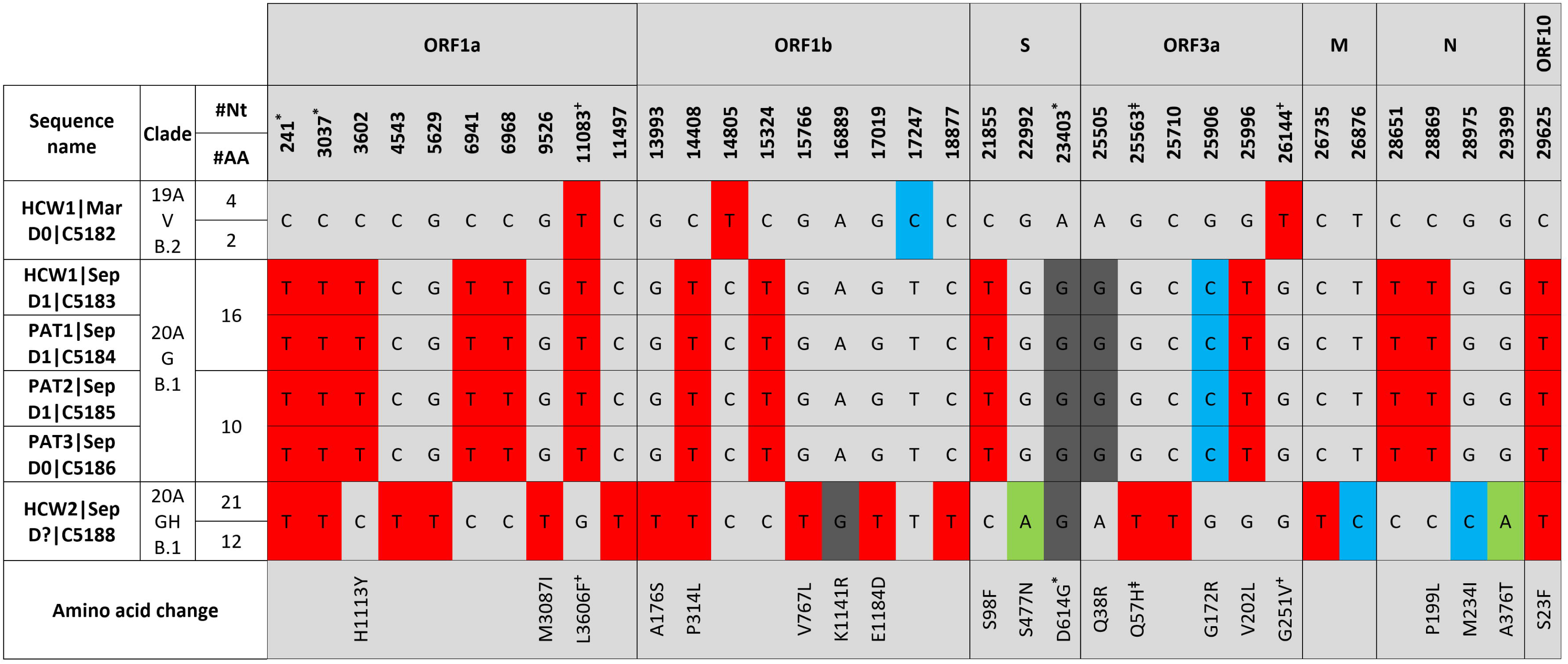
Nucleotide and amino acid comparison of the nosocomial SARS-CoV-2 genomes to the reference genome Wuhan-Hu-1. * Mutations defining GISAID’s G clade; + Mutations defining GISAID’s V clade; L Mutation defining GISAID’s GH clade, # Nt = number of nucleotide changes compared to Wuhan-Hu-1; # AA = number of amino acid changes compared to Wuhan-Hu-1; Clade = respectively Nextstrain (19A; 20A); GISAID (V; G; GH); and Pangolin (B.1; B.2) nomenclature; PAT = patient; HCW = health care worker; ORF = open reading frame; M = membrane; S = Spike; N = Nucleocapsid; D = days post symptom onset / diagnosis; Mar = March; Sep = September

### Assessment of SARS-CoV-2 specific humoral response and neutralizing antibodies

We then measured the presence of total SARS-CoV-2 specific antibodies as well as individual IgA, IgM, and IgG titers in the serum of the 4 individuals in the transmission cluster using all available samples. As antibody kinetics can depend on the antigen [20], we measured responses against the N protein and the RBD of the Spike protein. In addition, we performed *in vitro* viral neutralization tests on serum and swab samples to determine whether these antibodies have neutralizing capacity. HCW1 clearly seroconverted because 3 months after primary infection, she still displayed high serum IgG titers as well as neutralizing antibodies (200 NT50) at levels higher than those of the 3 patients 2 weeks post diagnosis (Table 1). As expected, serum IgM and IgA could not be detected at this timepoint. Since antibody testing was not available during the first wave of the pandemic in Belgium, and no baseline sample was taken upon re-infection, we could not evaluate whether her neutralization titers had been higher closer to primary infection or disappeared closer to re-infection. However, during re-infection, a rapid rise in neutralizing antibodies could be observed within 7 days pso (1309 NT50) which further increased 14 days later (>1600 NT50) in line with the high IgG titers and somewhat lower IgA and IgM titers at these timepoints (Table 1). PAT2 showed full seroconversion 13 days post symptom onset (pso) and displayed high IgA, IgM and IgG titers as well as a low neutralizing antibody response (75 NT50). For PAT1 however, who was first to develop symptoms, IgM levels were below the cut-off 17 days pso although IgA and IgG were clearly present as well as neutralizing antibodies (100 NT50). Finally, for PAT3 only low IgA titers and very low levels of neutralizing antibodies (<50 NT50) were detected 12 days post diagnosis which corresponds with his asymptomatic disease and high Ct values upon diagnosis (Table 1). No neutralizing antibodies were detected in any of the nasopharyngeal swabs including those of HCW1 taken at day 0 and day 7 pso.

### Potential onward transmission from the re-infection case

How transmission exactly occurred within this cluster of 4 individuals as well as its origin remain unclear. The identical genomes and timelines would suggest one index patient who infected the others. All 3 patients tested negative for SARS-CoV-2 upon admission to the hospital with a diabetic foot problem 4 to 6 weeks prior to this outbreak (Table 1). They were put on compulsory bed rest, stayed in private rooms, and only received a maximum of 1 visitor a day as per hospital rules. It is unlikely that HCW1 contracted the virus and transmitted it to the 3 patients as HCW1 developed symptoms 4 days after PAT1 and did not nurse PAT1. One explanation could be that an asymptomatic visitor of PAT1 infected both PAT1 and HCW1 who then transmitted the virus while nursing PAT2 and PAT3. However, none of the close contacts of HCW1, PAT1, and PAT3 tested positive for SARS-CoV-2, although cases could have been missed. PAT2 reported symptomatic SARS-CoV-2 infection in one of her relatives 2 days after her diagnosis, but is unlikely to be the index patient (and rather infected the visitor) as PAT2 developed symptoms 3 days after PAT1 who she did not have contact with. Finally, nasopharyngeal swabs taken from HCW1 at diagnosis and 6 days later showed high viral loads (Avg Ct 19 and 25 respectively), but lower than at diagnosis of primary infection (Avg Ct 13), and contained replicating virus as indicated by RT-qPCR for negative strand RNA (Avg Ct 25.5 and 28.5 respectively). Yet, we were unable to culture virus from these swabs, but this might be because we had to dilute the otherwise cytotoxic swab medium to a level where virus isolation can fail.

## Discussion

One of the key questions in understanding SARS-CoV-2 immunity and predicting the course of the pandemic is for how long and how frequently primary infection protects against re-infection. Eight cases of re-infection have now been described showing intervals between episodes from 48 to 142 days. In this study, we describe another genuine case of SARS-CoV-2 re-infection with an interval of 185 days. The viral genomes from the first and second episode belonged to different SARS-CoV-2 clades and phylogenomic analysis showed that the closest relatives to these 2 genomes were strains collected mostly from Belgium around the same period.

We then quantified the humoral immune response in HCW1 after her first and second SARS-CoV-2 episode as it was suggested that asymptomatic and mild primary infections might not protect against re-infection. The fact that HCW1 fully seroconverted and even had significant levels of serum neutralizing antibodies 3 months after primary infection, suggests that she wasn’t an exceptional patient unable to mount a humoral immune response. The re-infecting virus also didn’t harbor any known Spike mutations that could have enabled escape from neutralizing antibodies induced during primary infection. The durability of the humoral immune response on the other hand, remains a debated issue. Although some early studies reported a rapid loss of humoral immunity [14,30] within 2-3 months in up to 40% of patients with mild disease [18], more recent evidence shows that neutralizing antibodies and IgGs actually reach a stable nadir after an initial decline [20,31], which persists for at least 5 to 7 months, presumably as short-lived plasma cells are replaced with long-lived antibody secreting cells [20]. However, as no blood sample was taken from HCW1 right before or at day 0 of the second episode, we could not reliably determine whether neutralizing antibodies persisted or whether a further loss allowed for the re-infection. Of note, neutralizing antibodies are only a marker of immunity and the antibody level needed to confer protection to SARS-CoV-2 is currently unknown.

The slightly milder clinical disease course of HCW1’s secondary infection with normal physiological parameters, is likely the result of the patient’s adaptive immunity being primed by the first infection. This corresponds with the lower viral loads and the strong and fast rise in serum IgG, and neutralizing antibody responses observed after re-infection while IgA and IgM levels remained rather low. Unfortunately, we were unable to compare these antibody levels to similar time points after the first episode.

It seems likely that HCW1 played a role in the spread of this outbreak as she provides the only link between some of the patients. Furthermore, although virus culture remained inconclusive, HCW1’s nasopharyngeal swabs contained replicating virus but no neutralizing antibodies which suggests the re-infecting virus was fully capable of onward transmission. However, none of her contacts tested positive, which might be reassuring.

In conclusion, we describe a case of SARS-CoV-2 re-infection in a young, immunocompetent patient who, in contrast to the Hong Kong case, developed an effective humoral response after primary infection. If cases like this substantially increase over the next few months, long-lived protective immunity against SARS-CoV-2 will not be likely, which would be in line with other human coronaviruses, and might impact current vaccine development strategies which are based on eliciting neutralizing antibody responses.

## Data Availability

All raw data are available through corresponding author or presented in the manuscript

## Acknowledgements

PS and JM are members of the Institute of Tropical Medicine’s Outbreak Research Team which is financially supported by the Department of Economy, Science and Innovation (EWI) of the Flemish government. Part of the research was covered by the Flemish Research Foundation [grant number FWO-G0G4220N to KA]. We thank Odin Goovaerts and Wim Adriaensen for carefully reading the manuscript as well as all sequence contributors to the GISAID database.

## Figure Captions

**Supplementary figure 1. Phylogenomic analysis of 6 genomes recovered from 3 patients (PAT) and 2 health care workers (HCW) within a panel of most closely related SARS-CoV-2 sequences**. A maximum likelihood phylogenetic tree (TIM2+F+I model) was inferred with IQ-TREE v1.6.12 using 500 nonparametric bootstraps and the -czb option to collapse near zero branch length and rooted using Wuhan-Hu-1 (MN908947.3) as outgroup. Leaf labels contain the country of origin, sequence name, GISAID accession ID, date, and region. The genomes of the first and second infection of HCW1, patients, and HCW2 are depicted in respectively red, blue and green text. The scale bar represents the average number of nucleotide substitutions per site.

