## Supplementary figures and images for "Symptomatic SARS-CoV-2 re-infection of a health care worker in a Belgian nosocomial outbreak despite primary neutralizing antibody response"

### Supplementary_figure_1

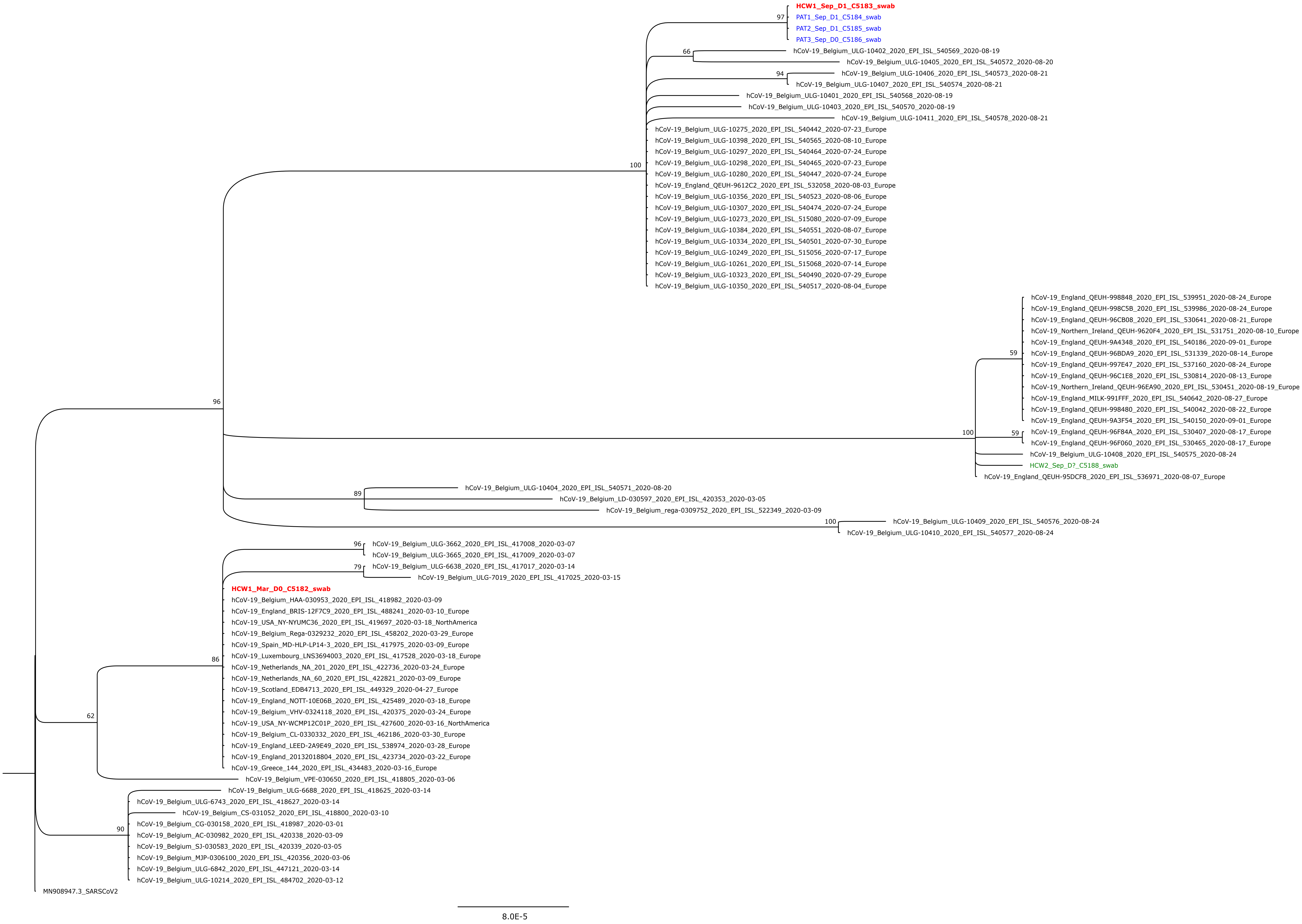
