## Supplementary_table_1 for "Symptomatic SARS-CoV-2 re-infection of a health care worker in a Belgian nosocomial outbreak despite primary neutralizing antibody response"

**Supplementary table 1:** hematological and biochemical parameters during HCW1’s primary and secondary SARS-CoV-2 episode.

|  | Episode 1 | | Episode 2 | |
| --- | --- | --- | --- | --- |
|  | **D1** | **D5** | **D7** | **D18** |
| Temperature (36.0 – 38°C) |  | 37.3 |  | 37.3 |
| Saturation (95 - 100%) | 100 | 98 |  | 99 |
| Respiratory rate  (12 - 20 breaths/min) | 12-20 | 12-20 |  | 12-20 |
| PO2  (83 - 108 mmHg) | 105 | 87.8 |  |  |
| PCO2  (32 – 45 mmHg) | **21** | 33.3 |  |  |
| Hemoglobin  (11.6 - 14.4 g/dL) | 14.6 | 13.7 | 14.2 | 13.5 |
| Trombocytes  (166 - 396 x 10E9/L) | 158 | **119** | 250 | 231 |
| WBC (4.2 - 10.3 x 10E9/L) | **3.22** | **3.41** | 7.75 | 6.12 |
| Absolute neutrophils (2.0 - 6.7 x 10E9/L) | **1.34** | **1.52** | 3.38 | 2.62 |
| Absolute lymphocytes (0.9 - 3.4 x 10E9/L) | 1.58 | 1.71 | 3.94 | 2.97 |
| Absolute monocyten (0.3 - 0.8 x 10E9/L) | 0.28 | **0.15** | 0.39 | 0.49 |
| Absolute eosinophils (0.02 - 0.25 x 10E9/L) | 0.01 | 0 | 0.03 | 0.02 |
| Absolute basophils (0.01 - 0.09 x 10E9/L) | 0 | 0 | 0.01 | 0.01 |
| APTT (23.0 - 31.0 sec) | 24.9 | 25.3 |  | 21.7 |
| PT  (78.0 - 123%) |  | **>130** |  | 122 |
| Creatinine (0.50 - 0.80 mg/dL) | 0.67 | 0.63 |  | 0.62 |
| CRP (<10 mg/L) | **13.7** | **27.1** | 7.2 | <4.0 |
| LDH  (120 - 246 U/L) | 159 | 200 | 168 | 137 |
| CK  (34 - 145 U/L) | 73 | 197 |  | 78 |
| Ferritine  (10 - 291 µg/L) |  |  | 52 |  |

D = day post symptom onset; PO2 = partial O_2_ pressure; PCO2 = partial CO_2_ pressure; WBC = white blood cells; APTT = activated partial tromboplastin time; PT = prothrombin time; CRP = C-reactive protein; LDH = lactate dehydrogenase; CK = creatine kinase. The reference range for each of the parameters is given between brackets and values that deviate are depicted in red.
