## Supplementary_table_2 for "Symptomatic SARS-CoV-2 re-infection of a health care worker in a Belgian nosocomial outbreak despite primary neutralizing antibody response"

**Supplementary table 2.** Ct values and sequencing statistics on nasopharyngeal samples taken from 3 patients and two health care workers

| Reference number | Patient ID | Collection | Diagnostic Ct | | ITM Ct | Genome  Coverage | Mean depth |
| --- | --- | --- | --- | --- | --- | --- | --- |
|  |  |  | E gene | N2 gene | E gene |  |  |
| C5182 | HCW1 | Mar_D0 | 15 | ND | 11.2 | 99.6% | 1148x |
| C5183 | HCW1 | Sep_D1 | 17.4 | 20.5 | 19.5 | 99.6% | 1024x |
| C5187 | HCW1 | Sep_D7 | 24.2 | 26.6 | 22.7 | 99.6% | 853x |
| C5184 | PAT1 | Sep_D1 | 13.1 | 15.9 | 13.5 | 99.6% | 803x |
| C5185 | PAT2 | Sep_D1 | 20.4 | 22.6 | 21.1 | 99.6% | 802x |
| C5186 | PAT3 | Sep_D0 | 28.1 | 31.7 | 27.3 | 99.6% | 269x |
| C5188 | HCW2 | Sep_D? | 29.0 | 31.5 | 28.6 | 99.6% | 499x |

ITM = Institute of Tropical Medicine; HCW = health care worker; PAT = patient; Ct = RT-qPCR cycle threshold; D = days post symptom onset / diagnosis; Mar = March; Sep = September
